# Development and Validation of an Eating Disorder Screening Tool for Children in Primary Care

**DOI:** 10.1101/2025.03.20.25324321

**Authors:** Dori Steinberg, Nickolas Jones, Megan Whelen, David Freestone, Jessica H. Baker, Lauren Davis, Megan Hellner, Madelyn Johnson, Taylor Perry, Bek Urban, Cara Bohon

**Affiliations:** Equip Health, Inc., Carlsbad, CA, United States of America; Steinberg Research Consulting; Stanford University

## Abstract

Clinical practice guidelines for medical professionals working with children and adolescents recommend routine screening for eating disorders. However, no validated screening tools exist for this age group. The aim of this study was to design and validate a brief, caregiver-based screening tool for eating disorders in children ages 6-12 that can be used in primary care. Caregivers of children aged 6-12 completed a battery of questionnaires (N=1,760). A subsample of caregivers completed semi-structured diagnostic interviews (n=255) to evaluate child eating disorder diagnosis. Standard data science practices were used to determine which questions were best at detecting an eating disorder and the most appropriate scoring threshold. Our final screener consists of two items that were effective at discriminating between children with an eating disorder and those without one and that were general enough to capture multiple diagnoses. As is recommended by organizations such as the AAP, this tool would ideally be administered to all caregivers of children ages 6-12 at yearly wellness visits beginning at the age of 6.

## Introduction

Eating disorders are life-threatening psychiatric conditions characterized by disturbances in eating or related behaviors that cause short and long-term impairment in physical and psychosocial functioning.^1^ Eating disorders are seen in children as young as 6, especially avoidant restrictive food intake disorder (ARFID).^2,3^ Rates of eating disorders among youth have nearly doubled over the past several years, with global estimates showing that 23% of children and adolescents engage in disordered eating (e.g., dieting, inappropriate compensatory behaviors).^4^ Despite this steep increase in rates of eating disorders and related behaviors, they are often missed in childhood due to a lack of available screening resources. Currently, it is estimated that the lag between onset of symptoms and treatment can be up to 7 years.^5^

Pediatricians are well positioned to identify early signs and symptoms of eating disorders due to frequent and prolonged engagement with children and their families.^6^ Current practices already include scheduled screenings for mental health disorders during wellness visits (e.g., autism, ADHD), and the American Academy of Pediatrics (AAP) recommends screening for eating disorders during the annual visit or sports exams. However, The US Preventive Service Task Force deemed evidence on screening in individuals ages 10 and older to be insufficient,^6^ indicating an overall need for more evidence on how best to screen for eating disorders. Given that eating disorder risk factors can start as early as age 6, there is a dire need for early eating disorder screening to interrupt disease progression.^7^

Several screening instruments exist to detect eating disorders in adults;^8–10^ however, there are no validated instruments for children.^6^ Moreover, existing screening tools were generally developed without inclusion of indicators of ARFID and were designed to detect an eating disorder diagnosis, not necessarily the behaviors that *precede* it. An effective screening tool should alert physicians of risk in presumptively healthy patients.^11^ As such, a useful childhood screener for eating disorders should alert pediatricians to problematic symptoms or risk before the onset of an eating disorder diagnosis.^12^ This allows early intervention to be put in place before symptoms worsen and, in turn, may stop the progression of the disease.

This study aimed to develop and validate an eating disorder screening tool for children for use in pediatric settings that can detect early signals of an eating disorder.

## METHODS

### Study Population and Design

Participants were recruited via CloudResearch Connect, an online platform that connects researchers with potential participants.^13^ There are millions of potential respondents engaged with the platform allowing access to research study participants that would not otherwise be available for study participation.^14^ For example, in comparison to clinic-based studies, bound by geography near the clinic location, we were able to recruit from a pool of participants across the U.S. and reach participants not a patient at a recruitment site, as well as harder to reach populations such as caregivers/parents^15^ whose family responsibilities may conflict with the ability to participate in an in-person research study or those with financial barriers that do not allow for regular clinic visits. This allows for a participant sample that better aligns with the study population of interest. Indeed, the CloudResearch Connect platform demographics closely align with that of the 2024 U.S. Census.^16,17^

Adult caregivers with a child between the ages of 6-12 were eligible. There were no other inclusion/exclusion criteria. Potential participants were provided a description of the study through the web-based platform; described as a study about the participants’ child’s eating behaviors. The study included two parts: 1) completing an online survey; 2) a virtual diagnostic interview. All survey completers were contacted approximately 1-week after survey completion with the opportunity to complete the interview. Participants who completed the survey were compensated $4.00 for their time. Participants who completed the interview were compensated an additional $11.00. Compensation amount is based on the CloudResearch Connect recommendation of at least $7.50/hour with a majority of studies offering incentives of $10/hour. The median survey completion time was 14-minutes and interviews ranged from 15-30 minutes. The study was reviewed and approved by the BRANY Institutional Review Board (New York, NY). Participants (caregivers) provided informed consent prior to study participation.

### Data Sources

All data were obtained from parents/caregivers. Children were not included in the study.

### Pediatric Eating Disorder Screener (PEDS) Scale Development

Clinicians and researchers with extensive experience in eating disorders and those with lived experience with an eating disorder developed a set of 12 initial questions to target transdiagnostic eating disorder symptoms that present in children ages 6-12 (Table 1). These questions were based on clinical presentations of eating disorders in primary care and currently available assessments of parent-based eating disorder assessment measures. We aimed to make the questions simple and not double-barreled. The full survey had good internal consistency (Cronbach’s ɑ = 0.82 [0.80, 0.84]). Participants completed the full PEDS as part of the initial survey battery and those that completed the survey were contacted on average 41 (SD=22) days after survey completion to evaluate test-rest reliability. The retest survey also had good internal consistency (Cronbach’s ɑ = 0.86 [0.84, 0.87]).

**Table 1.**
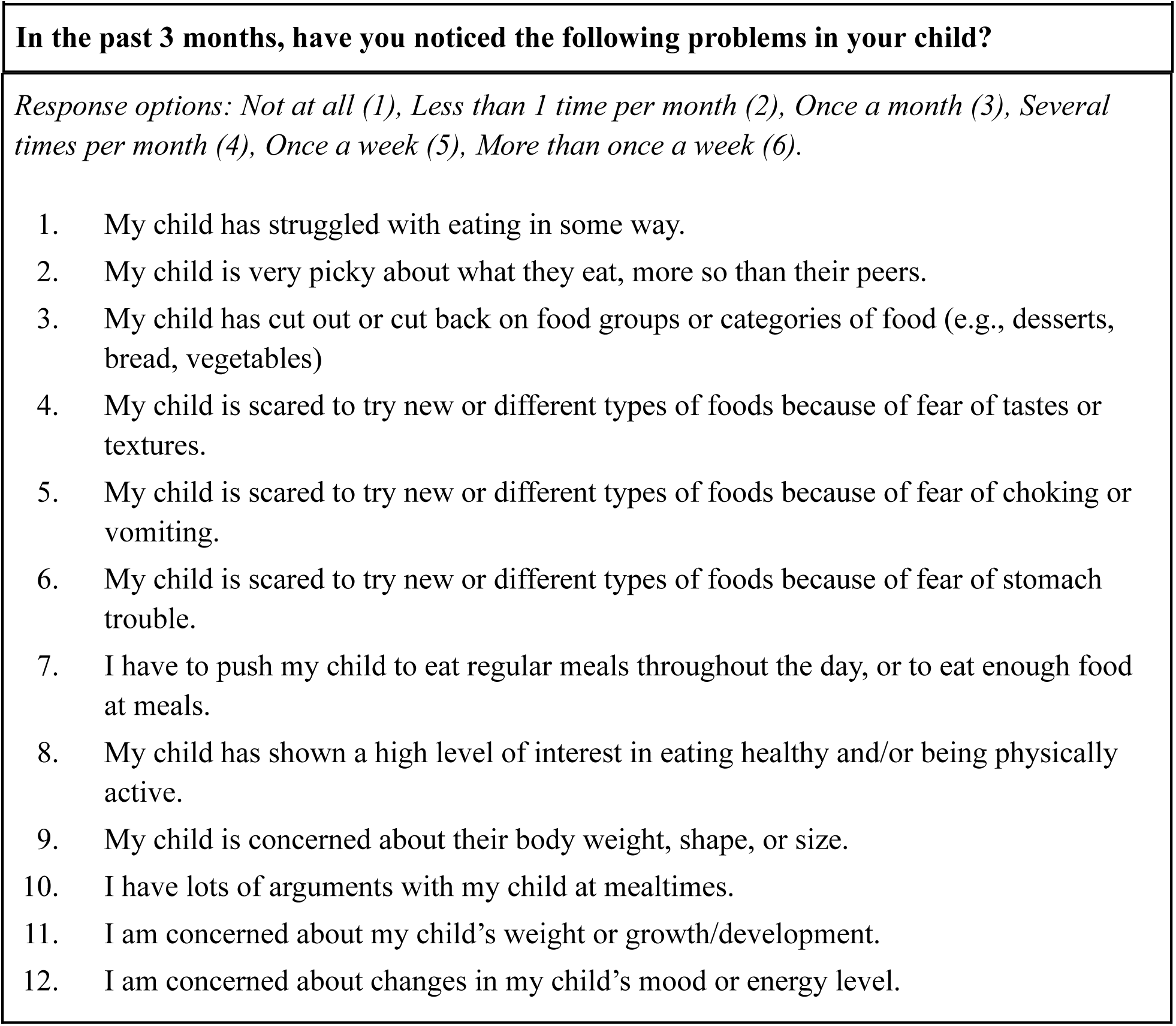
Initial set of pediatric eating disorder screening items for caregivers of children ages 6-12.

### Diagnostic Interview

Eating disorder diagnosis was determined using the Diagnostic Interview for Anxiety, Mood, and OCD and Related Neuropsychiatric Disorders-Child and Adolescent Version (DIAMOND-KID).^18^ The DIAMOND-KID is a semi-structured diagnostic interview for DSM-5 diagnoses in children ages 6-18 years old, and can be used with caregiver(s). We used the eating disorder module, which includes anorexia nervosa (AN), bulimia nervosa (BN), binge-eating disorder (BED), avoidant/restrictive feeding intake disorder (ARFID), and other specified and feeding disorder (OSFED). The DIAMOND-KID was chosen because of its excellent convergent validity and test-retest reliability,^18^ availability of a parent version, and inclusion of all eating disorders (e.g., ARFID, OSFED). All interviewers were masked to the participants’ survey responses.

Interviewers were psychologists or clinical psychology doctoral candidates with at least 5 years of experience completing semi-structured interviews and diagnostic assessments. All interviewers completed a 3-hour training course provided by the interview developers that trains raters to criterion and calculates the inter-rater reliability with the original interviewer. A reliability of at least .89 was required to ‘pass’ the course. Interviewers then completed role plays and were observed completing an interview prior to signing off on training. Interviewers also completed synchronous and asynchronous training sessions on the study protocol. Interviewers attended weekly supervision sessions, supervised by a clinical psychologist with over 10 years of experience in conducting eating disorder diagnostic interviews (JB) and a registered dietitian with 20 years of experience providing treatment for children and families with an eating disorder (MH).

As part of the interview process, interviewers classified some cases as “monitor,” when the child did not meet criteria for an eating disorder diagnosis, but caregivers reported symptoms/behaviors that were of concern (e.g., extreme food restriction but no weight loss, infrequent binge episodes, inappropriate levels of exercise for weight loss or muscle gain etc.). In these cases, even though diagnostic criteria were not met, there was general consensus among the interview team that more frequent monitoring would be warranted. Children identified as monitor cases may greatly benefit from early screening, as providers can intervene before behaviors progress/worsen. We depict the performance of monitor cases on our screener in Figure 2a. Otherwise, for all analyses, monitor cases are treated as having no diagnosis.

### Secondary Measures

ARFID symptoms were evaluated with the Pica, ARFID, Rumination Disorder Interview (ARFID subscale) Questionnaire - parent version (PARDI-AR-Q-PV),^19^ which is a 32-item caregiver report of ARFID symptoms in children ages 4 and up. The PARDI-AR-Q-PV includes symptom subscales and a binary diagnostic score that indicates whether the child likely meets DSM-5 criteria for ARFID.^19^

The Eating Disorder Examination Questionnaire Parent Version (EDE-Q-PV) evaluated symptoms related to anorexia nervosa (AN), bulimia nervosa (BN), and binge eating disorder (BED). The EDE-Q-PV is an adapted version of the 28-item EDE-Q where parents answer questions about their child’s eating disorder cognitions and behaviors in the past 28 days.^20^ The EDE-Q-PV has good internal consistency and correlates with the self-report version of the EDE-Q subscales.^20^ Parents report how often their child has experienced symptoms on a 7-point likert-type scale ranging from 0 (*no days*) to 6 (*every day*). The EDE-Q-PV consists of symptom subscales and a global score.^20^ A global score of 2.8 or higher is considered clinically significant.^21^

We included two additional eating disorder screening tools for comparison that were not developed for children but have been used with youth.^22,23^ Items were modified to change self-report to “my child.” The SCOFF consists of five questions addressing the core features of AN and BN;^8^ a score of 2 indicates a likely diagnosis of AN or BN. The Eating Disorder Screen for Primary Care (ESP) is a five-item screener developed for adults in primary care. Two or more items endorsed indicate a possible eating disorder.^24^

### Data Procedures and Statistical Analysis

Multilevel Bayesian ordinal regressions were used to characterize participant responses to the 12-item PEDS survey. The model included an intercept term and the following as random effects: the survey item; patient gender and age; DIAMOND-KID diagnosis, and interactions (diagnosis x item, diagnosis x gender, diagnosis x age, item x gender, item x age); and finally, a random intercept by participant identification number. To determine the most useful PEDS items for distinguishing cases that should be assessed further, we evaluated the discrimination score - the difference between the proportion of children with and without an eating disorder whose parent/caregiver responded with the top response option (‘*More than once a week*’) to each item. To balance screener length and function, we 1) trimmed the PEDS based on discriminability scores and inter-item correlations (retaining a single, clinically important item if it correlated highly with another item) and 2) evaluated the extent to which including more items reasonably increased the PEDS efficacy in flagging cases that should be assessed further.

All analyses were conducted using R version 4.4.1 (R Core Team, 2022) and tidyverse version 2.0.0.^25^ Model fitting was done using the brms package version 2.22.0.^26,27^ Targets version 1.8.0 was used for project management.^28^

## Results

### Participant Characteristics

The study was advertised to 1,925 caregivers, 1,760 completed the survey battery, and 729 completed the retest. Based on the general prevalence of eating disorders in the population of 10%, we aimed to complete at least 200 caregiver interviews. All caregivers that completed the survey were invited to complete the diagnostic interview, and 255 completed the interview. We compared the demographic characteristics of participants who completed the interview and those who did not and found no statistically significant differences. Parents/caregivers who completed the interview were, on average, age 39.4 (SD = 6.5), primarily White (72%), cisgender women (66%) and married (75%) (Table 2). Nearly a third of children whose caregivers participated in the interview were diagnosed with an eating disorder on the DIAMOND-KID (n = 82; 32%). Diagnoses included ARFID (n = 48; 59%), OSFED (n = 26; 32%), BED (n = 3; 4%), BN (n = 3; 4%), and AN (n = 2; 2%). Within OSFED, there were 10 cases of atypical AN, 2 cases of low-frequency BED, and 14 cases that fell into the “other” category, 12 of which had ARFID-like features. Nineteen children were identified as a “monitor” case by caregiver report to interviewers.

**Table 2.** Demographic characteristics of caregivers completing the interview and their children (N = 255)

| **Variable** | **Child**<br>***M (SD)*** | **Caregiver**<br>***M (SD)*** |
| --- | --- | --- |
| **Participants** | - | 255 |
| **Age** | 9.3 (1.9) | 39.4 (6.5) |
|  | **N (%)** | **N (%)** |
| **Gender** |  |  |
| Cisgender women/girl | 115 (45%) | 169 (66%) |
| Cisgender men/boy | 138 (54%) | 82 (32%) |
| Nonbinary/agender/gender fluid | 1 (0%) | 2 (1%) |
| Did not report | 1 (0%) | 2 (1%) |
| **Race** |  |  |
| White | 178 (70%) | 183 (72%) |
| Asian | 11 (4%) | 13 (5%) |
| Black/African American | 42 (17%) | 39 (15%) |
| Native American | 1 (0%) | 1 (0%) |
| Other, multiracial/biracial | 18 (7%) | 12 (5%) |
| Did not report | 5 (2%) | 7 (3%) |
| **Ethnicity** |  |  |
| Hispanic | 35 (14%) | 24 (9%) |
| Non-Hispanic | 214 (84%) | 223 (88%) |
| Did not report | 6 (2%) | 8 (3%) |
| **Previously diagnosed with ED by medical professional** |  |  |
| Yes | 11 (4%) | 20 (8%) |
| No | 240 (94%) | 229 (90%) |
| Unsure | 2 (1%) | 3 (1%) |
| Did not report | 2 (1%) | 3 (1%) |
| **Marital Status** |  |  |
| Married or long-term partner | - | 192 (75%) |
| Divorced | - | 33 (13%) |
| Never married | - | 29 (11%) |
| Did not report | - | 1 (0%) |
| **Income** |  |  |
| \$0 - \$50,000 | - | 66 (26%) |
| \$51,000 - \$90,000 | - | 70 (28%) |
| \$91,000 - \$130,000 | - | 55 (22%) |
| More than \$130,000 | - | 59 (23%) |
| Did not report | - | 4 (2%) |

### Item Selection for Pediatric Screener

Figure 1 shows how caregivers answered each of the 12 items on the PEDS. Items are sorted by discriminability (read left to right, top to bottom). Overall, the responses on some items indicated discrimination between children with and without an eating disorder. Items 1, 2, 3, 4, and 7 showed the greatest ability to discriminate between presence and absence of a diagnosis.

**Figure 1.**
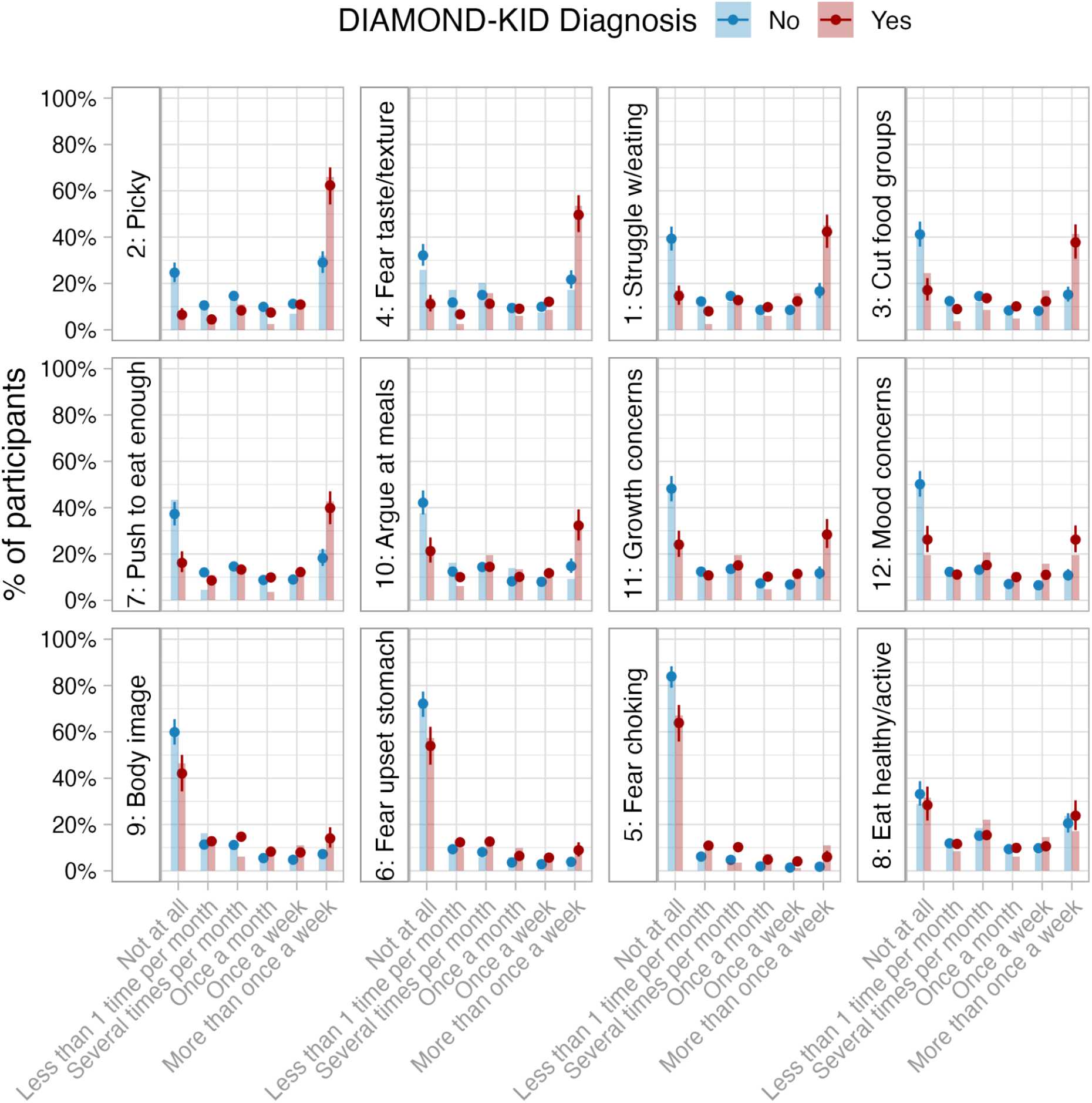
Within each panel, the x-axis gives the possible response options, and the y-axis represents the proportion of caregivers who gave that response. In red are the children who were subsequently diagnosed with an eating disorder during the DIAMOND-KID interview with the caregiver, those in blue were not. The bars show the raw proportions, the circles and error bars show the results of the ordinal regression model. Items are ordered by discriminability, that is, the extent to which the top box discriminates between those with and without a diagnosis on the DIAMOND-KID.

Next, we evaluated the performance of various combinations of items 1, 2, 3, 4, and 7. For each possible combination, we calculated the estimated proportion of children who would screen positive. We chose *a priori* to prioritize true positives in order to decrease the probability of false negatives. Further, the goal of the screener is not to only identify those children with a current eating disorder diagnosis, but also those at risk. We examined each possible screener’s performance in the full sample and by gender and diagnosis (defined as ARFID vs non-ARFID). We tested 2, 3, and 4-item screeners and performance was highly similar. Therefore, we opted for a 2-item screener to maximize simplicity of administration. Since items 1, 2, and 4 individually have the highest discriminability (Figure 1), we compared items 1 and 2 against items 1 and 4. While they performed similarly, the combination of items 1 (“My child has struggled with eating in some way”) and 2 (“My child is very picky about what they eat, more so than their peers”) had a slightly greater ability to detect non-ARFID diagnoses (at a threshold of 5 or greater, ARFID: 83% (74%, 91%); Non-ARFID: 76% (59%, 89%)), so these items were retained for the final screener.

To further simplify use for clinic settings, we evaluated various response thresholds for the final items (i.e., items 1 and 2). For the choice of threshold, monitor cases were included to evaluate how well the threshold identifies this “at risk” group. Figure 2a (top left) shows how the choice of threshold determines how many children would screen positive and how many of those would likely have an eating disorder (red line; those without an eating disorder are in blue). As the threshold cutoff increases, the number of children who screen positive decreases, but the discriminability between those with and without an eating disorder also increases. At a threshold of 5 or greater on either item, 48% of children who do not have an eating disorder screen positive (boys: 43% [CI: 33%, 53%]; girls: 52% [CI: 42%, 63%]). These children may still benefit from a positive screen that leads to further evaluation, given the frequency of their eating difficulties.

**Figure 2.**
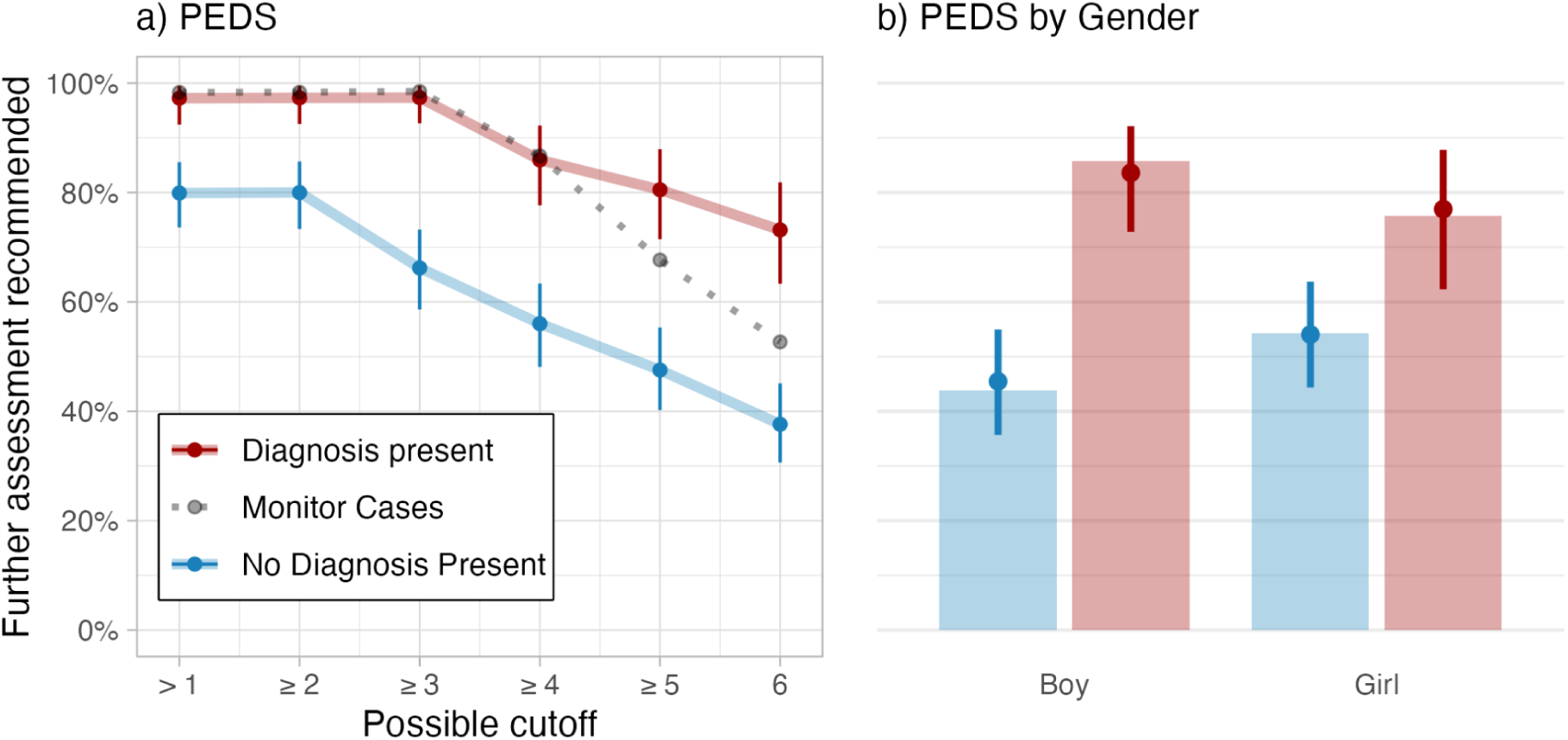
Performance of the pediatric screener by possible cutoff (panel a) and gender when the cutoff is 5 (panel b); *Note:* The figure in Panel *a* shows the percentage of children who would be flagged at different cutoffs for the final screener items (Items 1 and 2). Shows highest score for either item 1 or item 2.

On the other hand, at this threshold 81% of children with an eating disorder would screen positive (boys: 84% [CI: 73%, 92%]; girls: 77% [CI: 62%, 87%]). Using this threshold would also identify 68% of monitor cases (n = 19, gray dashed line in Figure 2a; those the clinician believed were at risk for an eating disorder). Thus, data suggests that a cutoff of 5 (*once a week*) on *either* screener item provides an indication of a disordered behavioral pattern (i.e., a high risk response). The choice of recommending an elevated score on *either* item maximizes the screener’s efficacy in flagging concerning cases given that one item is more general and the other is ARFID-specific (and more common in a pediatric population). At this threshold the final screener items yield sensitivity of .81 [.72, .88], a specificity of .50 [.43, .57], and area under the ROC curve of .66 [.61, .72]. However, we largely avoid interpreting the screener in terms of sensitivity and specificity because those metrics emphasize identifying children who have a disorder and excluding those who do not have one. However, there is a benefit in identifying children who are engaging in problematic behaviors that do not meet a diagnosis (e.g., intervening to prevent the development of a diagnosis).

The retest surveys were filled out a little over a month after the initial survey (M = 41, SD = 22 days). The intra-class correlations showed modest test-retest reliability (ICC = 0.68 [0.66, 0.71]) for the final two screener items. Notably, because of the Likert response options for the items and the timeframe for test-retest (on average, 41 days), it is possible that symptom frequencies may have truly shifted from the first to the second survey. On the Likert scale response options, participants gave a similar response, within one adjacent response option, 75% (SE = 2%) of the time on both surveys. Those with a positive screen on the first survey were 69% (2%) likely to also have a positive screen on their second. Those with a positive screen on their second survey were 85% (2%) likely to have had a positive screen on their first.

### Performance of Common Screeners

For illustrative purposes, we evaluated how well other commonly used eating screeners/surveys performed in our sample (Figure 3). The EDE-Q-PV performed poorly in our sample, as did the SCOFF. The ESP did reasonably well, flagging 62% of children with an eating disorder, and flagging 46% without. However, these surveys are not designed for a pediatric sample and do not include items assessing ARFID, the most common diagnosis in this pediatric age range. Finally, the PARDI-AR-Q was most effective at discriminating, flagging 54% of children with an eating disorder and only 17% without, but this is specific to an ARFID diagnosis only.

**Figure 3.**
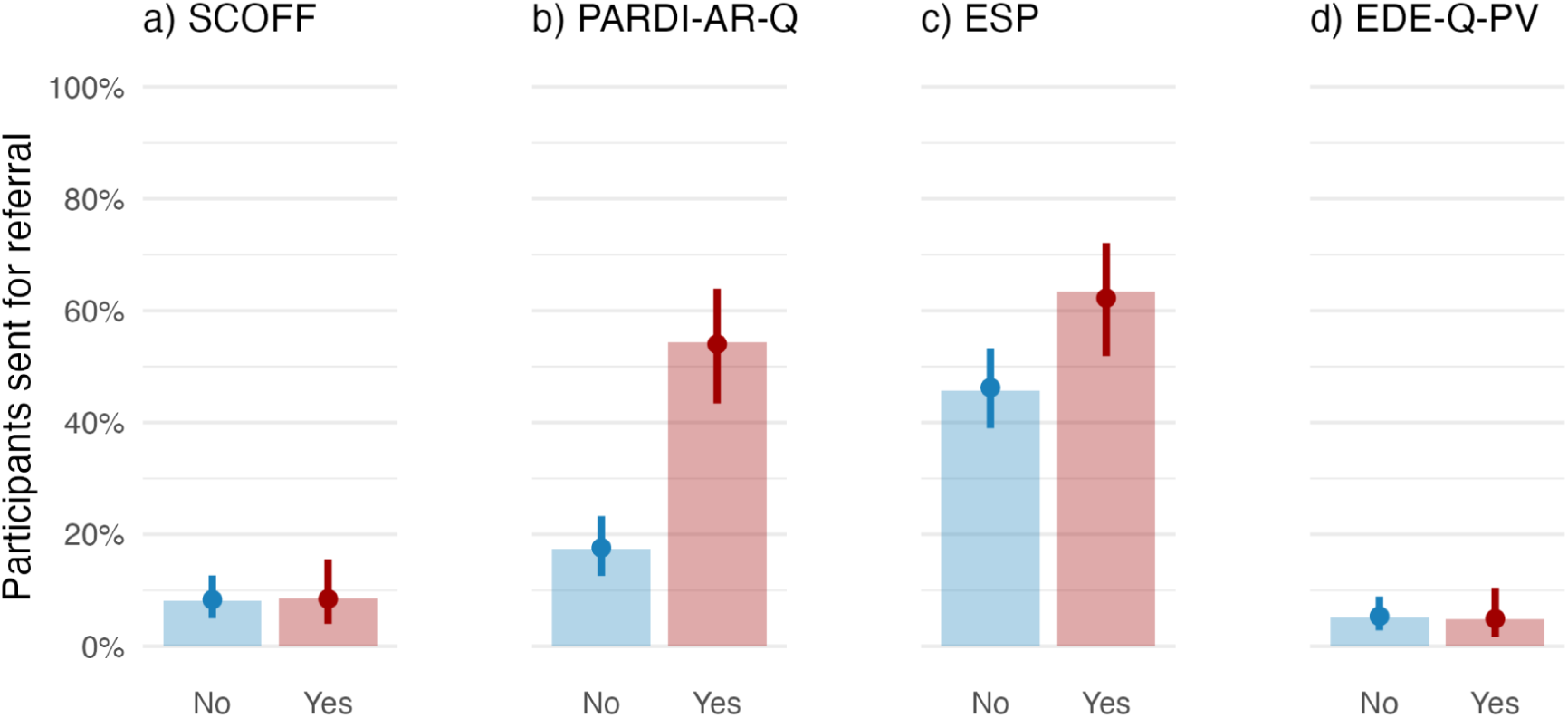
Proportion of participants who would screen positive on their performance on other screeners commonly used in the eating disorder field.

## Discussion

We present a brief and easy to use two-item screening instrument that can be used to screen for eating disorders and related behaviors that may onset prior to a diagnosis. Most childhood and adolescent health organizations recommend regular screening for eating disorders in children;^6,29–31^ however, a validated screening instrument for this age group does not exist.

The presented screener is brief and easily scored, an important consideration for implementation. A response indicating a frequency of once a week or greater on the item “My child has struggled with eating in some way” or on the item “My child is very picky about what they eat, more so than their peers” represents a reasonable cutoff for identifying high risk children in need of further assessment. The choice of recommending an elevated score on *either* item maximizes its efficacy in identifying possible ARFID and non-ARFID cases. The screener was specifically developed to be transdiagnostic and include symptoms relevant to all eating disorders, including ARFID and OSFED, given these particular diagnoses are more common in pediatric populations and have an earlier age of onset.^32,33^ Indeed, we showed screeners that do not include these symptoms (e.g., SCOFF) have a low response rate and do a poor job of discriminating those with an eating disorder from those without in this age group. Notably, the chosen cutoff for the screener is data-driven, yet patients who score lower than the cutoff may still need monitoring or be clinically appropriate to flag via clinical judgment.

Although we do not focus on sensitivity and specificity, the sensitivity–or proportion of positive cases screened that indeed have an eating disorder diagnosis–observed here is similar to other commonly used screeners in medical practices. For example, in a meta-analysis the PHQ-9 shows an overall sensitivity of .88,^34^ the GAD-2 sensitivity of .80,^35^ Vanderbilt ADHD diagnostic rating scale sensitivity .80,^36^ and the Modified Checklist for Autism in Toddlers Revised Follow-up sensitivity of .78.^37^ Although our specificity may be lower in comparison, the aim of the PEDS is to capture symptoms early to intervene before a diagnosis is present whereas many of the previously listed screens identify a probable diagnosis by replicating diagnostic criteria as a question. We placed priority on decreasing false positives, as a miss only extends the time lag from symptom onset to treatment, whereas a false negative results in minimal harm. As with any screener, providers will need to follow-up positive screens with evaluation of growth charts for observable deviations and further interviews with parents and children to determine the presence and severity of symptoms.

Despite strengths, this study is not without limitations. First, we obtained parent/caregiver report. Given eating disorders tend to be secretive, it is possible parents may not be aware of the presence or extent of some symptoms. However, caregivers are often the first to notice changes in eating behaviors and/or weight whereas young children may have difficulty understanding the impact and/or distress of such changes.^38,39^ It is common for parent report to be used for children, particularly when reporting on mental health concerns (e.g., autism, ADHD) that may be difficult for young children to understand and articulate. Additionally, this allows for change in behaviors to be evaluated more reliably across time as caregivers may have better recall of their child’s eating behavior across childhood and into early adolescence.

Second, prospective interview data was obtained approximately 1-week after survey completion. It is possible some children could have met criteria for an eating disorder at a later time. Third, we used the platform CloudResearch Connect to recruit participants, which may result in some selection bias (e.g., participants had to be part of the platform). However, in contrast to more traditional forms of study recruitment (e.g., social media, clinic-based recruitment) these platforms implement quality control and vetting of participants (e.g., low risk of bots), high respondent and follow-up rates,^13^ sample diversity similar to the U.S. census,^17,40^ and opportunities to reach a broader participant base–a major limitation of clinic-based studies. Moreover, research conducted in survey panels and population-based samples have similar results.^15,41^ The resulting sample for this study had racial and economic distributions similar to that of the U.S. population, increasing generalizability. Finally, it is possible children could screen positive due to another condition or upon further evaluation, an eating disorder is not present. However, the goal of this screener is <u>not</u> to identify eating disorder diagnoses. Children who screen positive may still benefit from more frequent monitoring and early intervention or identify other health concerns that were previously unrecognized. As with all screeners, it is imperative for the provider to follow-up with further evaluation and assessment to rule in or out the presence of an eating disorder or related behaviors.

## Conclusion

We created a brief, two-item screener to be used in pediatric practices to identify children *at risk* for an eating disorder. As is recommended by organizations such as the AAP, this tool would ideally be administered to all caregivers of children ages 6-12 at yearly wellness visits beginning at the age of 6. At the patient level, regular use of the instrument will allow for changes to be observed (e.g., moving from low to high risk), and at the population level, the routine detection of unrecognized cases. Although other screeners exist, the PEDS outperformed the other measures, is shorter, and has a less complicated scoring algorithm. Moreover, the PEDS specifically includes items relevant to ARFID, which are not explicitly addressed in other available screeners. Our next steps include developing a full implementation package for providers that includes the screener, scoring and recommendations for follow-up, and early intervention resources for use with patients and families.

## Data Availability

All data produced in the present study are available upon reasonable request to the authors.

